# Validation of gender identity and sexual behavior stigma measures among transgender women in the United States: Exploratory and confirmatory factor analyses

**DOI:** 10.64898/2026.11.17.26368277

**Authors:** Jason M. Lo Hog Tian, Kaitlyn Atkins, Sarah M. Murray, Jennifer L. Glick, Irah L. Lucas, Mariah Valentine-Graves, Savannah Winter, Joanna A. Caldwell, Travis Sanchez, Stefan Baral, John Mark Wiginton

## Abstract

**Introduction:** Transgender women experience stigma related to both gender identity and sexual behavior, yet robust measurement tools remain limited. This study evaluated the factor structure and external construct validity of the Measures of Attributional Stigma for gender identity (MAS-GI) and sexual behavior (MAS-SB) among transgender women in the United States.

**Methods:** We analyzed cross-sectional data from the Transgender Women’s Internet Survey and Testing (TWIST) platform collected from 2023–2025 (N=2,901). Participants completed parallel 14-item stigma measures attributed separately to gender identity and sexual behavior. The sample was split into exploratory factor analysis (EFA; n=1,951; 2023–2024) and confirmatory factor analysis (CFA; n=950; 2024–2025) samples. External construct validity was evaluated via associations with depression symptoms, posttraumatic stress disorder (PTSD), social capital, transgender identity pride, and healthcare utilization.

**Results:** EFA identified three MAS-GI factors (family stigma, anticipated healthcare stigma, and enacted social stigma) and two MAS-SB factors (global healthcare stigma and social/institutional stigma). CFA confirmed both structures with good to excellent model fit. All stigma dimensions were positively associated with depression and PTSD. Anticipated healthcare and enacted social stigma from the MAS-GI and both MAS-SB dimensions were negatively associated with social capital. Greater anticipated gender identity healthcare stigma associated with lower transgender identity pride. Several stigma dimensions were also positively associated with HIV/STI testing, PrEP use, and mental healthcare utilization.

**Conclusions:** The MAS-GI and MAS-SB demonstrate factorial validity and preliminary external construct validity among transgender women. These measures enable precise assessment of stigma by attribution and source and inform stigma surveillance and intervention evaluation.

## Introduction

Transgender women experience pervasive stigma stemming from multiple, interconnected systems of oppression including cissexism, transmisogyny, and heterosexism. Cissexism and transmisogyny produce stigma directed at gender identities and expressions that differ from cisgender norms, while heterosexism produces stigma directed at sexual behaviors, relationships, and identities that differ from what is perceived to be normative sexual behavior i.e., heterosexuality (1, 2). Gender identity stigma refers to the devaluation, discrimination, and negative treatment directed toward individuals whose gender identity differs from the sex assigned to them at birth within systems of cisnormativity, while sexual behavior stigma refers to negative treatment based on one’s sexual practices, partner characteristics, or perceived promiscuity, morality, or disease risk associated with certain sexual behaviors within systems of heteronormativity (3, 4). Given that many transgender women express a sexual orientation other than heterosexuality, transgender women can be subjected to both gender identity and sexual behavior stigma. Minority stress theory posits that stigma and discrimination constitute chronic stressors that contribute to adverse health outcomes among marginalized populations (5-7). Within this framework, understanding how stigma experiences are attributed to gender identity or sexual behavior can provide a better characterization of how these stigmatized attributes affect individuals and potential distinct relationships with key health outcomes.

The consequences of stigma for transgender women are well documented. Stigma has been associated with poor mental health outcomes including elevated rates of depression, anxiety, and post-traumatic stress disorder (8-10). Further, experiences of discrimination and violence have been linked to substance use, suicidal ideation, and suicide attempts in transgender populations (9, 11). Stigma also operates as a fundamental driver of HIV risk among transgender women, who bear a disproportionate burden of HIV infection globally (12, 13). Among transgender women, anticipated stigma in healthcare settings has been shown to lead to avoidance of HIV testing, prevention services, and treatment, while enacted stigma from healthcare providers has been associated with delayed care-seeking and disengagement from medical care (14-17).

Despite the recognized importance of stigma as a determinant of health in transgender communities, significant measurement gaps persist. Many studies examining stigma among transgender women have relied on measures developed for other populations, such as men who have sex with men or other cisgender sexual minority populations (18, 19). Some researchers have adapted existing stigma scales for use with transgender populations, and several transgender-specific stigma measures have been developed or adapted and validated for use (20-23). However, there are few measures that assess stigma as it relates to specific gender identity or sexual behavior. Attributional measures that assess the same stigma experiences across both domains can help identify whether specific sources or contexts of stigma are more strongly linked to one attribution versus the other, providing a greater understanding of how stigma affects transgender women who can often experience stigma related to both their gender identity and sexual behavior.

Wiginton, Maksut (22) adapted parallel 13-item measures of gender identity stigma and sexual behavior stigma for transgender populations using pilot data from the 2019 cycle of the Transgender Women’s Internet Survey and Testing (TWIST) study. In that exploratory analysis of 396 transgender women, the authors identified a two-factor structure for gender identity stigma (social stigma and institutional stigma/violence) retaining 12 items, and they identified a single-factor structure for sexual behavior stigma retaining nine items. The scale demonstrated preliminary evidence of associations with sexual health outcomes, suggesting evidence of construct validity. In subsequent years of TWIST, the scales were adjusted by removing consistently poor performing items and adding three new anticipated healthcare-related stigma items. While the initial work provided important preliminary evidence of factor structure and construct validity, psychometric exploration and validation of the updated scales with a larger, more geographically and racially/ethnically diverse sample is needed to confirm the factor structure, assess measurement stability, and provide stronger evidence of construct validity drawing on a range of constructs from the nomological network.

We refer to these instruments collectively as the Measures of Attributional Stigma (MAS) scales, which assess the same set of stigma experiences and distinguish them by attribution: stigma attributed to gender identity (MAS-GI) and stigma attributed to sexual behavior (MAS-SB). This study aims to (1) evaluate the factor structure and psychometric properties of the MAS-GI and MAS-SB scales among transgender women using exploratory factor analysis (EFA) and confirmatory factor analysis (CFA) in independent samples, and (2) examine preliminary external construct validity through associations with psychosocial and healthcare utilization outcomes.

## Methods

### Study Design

This study uses cross-sectional data (n=2,901) from the Transgender Women’s Internet Survey and Testing (TWIST) collected from December 16, 2023 to May 2025. Methods for the annual TWIST survey have been previously described (24). In brief, participants were recruited through online advertising including mobile dating apps and social media platforms to complete an online survey. To be eligible for the study, participants had to be ≥15 years old, identify as a transgender woman or transfeminine non-binary person (transgender hereafter), reside in the United States, and report ever having had oral, anal, or vaginal sex. Recruitment was monitored to improve representation across sociodemographic and geographic characteristics. To minimize duplicate and potentially fraudulent submissions, TWIST employed automated survey-quality screening, including CAPTCHA verification, survey completion checks, response consistency checks, and bot-detection measures. Flagged responses underwent additional manual review, including examination of IP address and response patterns, and only submissions meeting the study’s data-quality criteria were retained in the analytic dataset. Participants ≥18 years provided online written informed consent, and participants 15–17 years provided online informed assent (parental consent waived by the Emory University Institutional Review Board). Participants were not compensated and could withdraw from the study at any time. The original study was approved by the Institutional Review Board of Emory University.

### Measures

#### Stigma

Gender identity and sexual behavior stigma were assessed using the MAS scales, initially developed for use among men who have sex with men in sub-Saharan Africa and the United States, and subsequently adapted for use with transgender women in the United States (22, 25, 26). Scale adaptation was informed by input from a community advisory board comprising transgender women. The MAS scales assess the same 14 stigma experiences with parallel items attributed to gender identity (MAS-GI) and sexual behavior (MAS-SB). Participants could respond to each item with “No”, “Yes, but not in the last 12 months”, “Yes, in the last 12 months”, “I prefer not to answer”, and “Don’t know”. “I prefer not to answer” and “Don’t know” responses were coded as missing. To create subscale scores for the MAS-GI and MAS-SB, item responses were coded ordinally (0 = “No”, 1 = “Yes, but not in the past 12 months”, 2 = “Yes, in the past 12 months”) and subscale scores were calculated by summing the ordinal item responses with higher scores indicating a greater number and recency of sigma experiences. The scores were then standardized to facilitate interpretation. Unlike the prior validation study, which dichotomized affirmative responses due to small cell sizes (22), the larger sample in the current study permitted retention of the original ordinal response structure, allowing for more precise measurement and the use of polychoric correlations in factor analyses.

#### External construct validity measures

Depression was measured using the 9-item Patient Health Questionnaire (PHQ-9) (27), PTSD symptoms using an 8-item version of the PTSD Checklist for the DSM-5 (PCL-5) (28), social capital using 6 items adapted from Kerrigan, Telles (29), and transgender identity pride using the 8-item Pride subscale of the Transgender Identity Scale (20, 21). For all measures, items were summed with higher scores indicating greater levels of the construct. Participants who did not answer a majority of the items for a scale were counted as missing. Internal consistency of these scales was good with Cronbach’s alphas of 0.90 for depression, 0.91 for PTSD, 0.83 for social capital, and 0.88 for transgender pride.

Several healthcare engagement outcomes were also collected. Participants were asked if they had ever been tested for HIV, if they were tested for an STI in the past 12 months, if they had ever taken HIV pre-exposure prophylaxis (PrEP), if they had seen a healthcare provider about physical health in the past 12 months, and if they had seen a healthcare provider about mental health in the past 12 months. Responses were dichotomized as yes/no with “prefer not to answer” and “don’t know” responses being categorized as missing. Lifetime HIV testing and PrEP use were used due to low past-year incidence for both outcomes.

### Statistical Analysis

To support independent scale development and validation, the sample was divided by survey wave. The EFA sample (n=1,951) consisted of participants from the 2023–2024 TWIST survey cycle and the CFA sample (n=950) consisted of participants from the subsequent 2024–2025 cycle. In the 2024-2025 cycle, participants were randomized to answer certain sections of the survey including the MAS scales to reduce participant burden, hence the reduced sample size. Analyses were conducted separately for the MAS-GI and MAS-SB scales to establish the independent psychometric properties of each attribution-specific measure.

EFA was conducted to examine the underlying structure of the stigma items. Sampling adequacy was evaluated using the Kaiser–Meyer–Olkin (KMO) statistic and Bartlett’s test of sphericity (30, 31). A KMO ≥0.50 and a significant Bartlett’s test (p<0.05) indicates adequate sampling to detect underlying factors and data suitability for factor analysis. A principal components analysis (PCA) was then conducted on a polychoric correlation matrix given all included items were ordinal (32). A scree plot was then generated and parallel analysis performed. The number of factors to retain was determined using multiple criteria, including the result of the parallel analysis, number of eigenvalues >1, scree plot inspection (number of points above the “elbow”), and conceptual interpretability. The EFA was conducted, and factors were extracted using principal axis factoring with oblimin rotation, which allows factors to correlate while minimizing cross-loadings and is recommended when underlying constructs are expected to be related (33, 34). Items with low loadings (<0.40) across all factors and then with substantial cross-loadings (i.e., loaded ≥0.40 on a main factor but also ≥0.30 on a second factor, with a difference of ≤0.20 between the two loadings) were removed one-by-one and the EFA repeated until all issues were resolved (35). Ordinal alpha, which uses polychoric correlations to provide more accurate estimates for scales with ordinal items, was calculated to assess the internal consistency of each factor’s items (36, 37). Ordinal alpha was interpreted as ≥0.90 being excellent, 0.80–0.89 being good, 0.70–0.79 being acceptable, 0.69-0.6 being questionable, and <0.60 being poor (38).

The final factor structure identified in the EFA was evaluated using CFA in the 2024-2025 TWIST sample. Models were estimated using a diagonally weighted least squares estimator (DWLS/WLSMV) appropriate for ordinal indicators (39). Model fit was assessed using the Comparative Fit Index (CFI), Tucker–Lewis Index (TLI), Root Mean Square Error of Approximation (RMSEA), and the Standardized Root Mean Square Residual (SRMR). Model fit was interpreted using criteria of CFI and TLI ≥0.95, RMSEA ≤0.06, and SRMR ≤0.08 as indicative of good model fit (40).

External construct validity was evaluated by examining associations between stigma subscale scores and psychosocial and healthcare engagement outcomes in the CFA sample. Associations with continuous psychosocial measures (depression, posttraumatic stress symptoms, social capital, and transgender identity pride) were assessed using Pearson correlation coefficients. Associations with binary healthcare engagement outcomes were estimated using modified Poisson regression with robust variance estimation to obtain prevalence ratios (PRs) and 95% confidence intervals (41). Separate bivariate models were fit for each stigma subscale and outcome. Statistical significance was defined as p<0.05 for correlation coefficients or a 95% confidence interval that does not span 1 for PRs. All analyses were conducted in R version 4.5.1 (42).

## Results

### Participant characteristics

Table 1 shows participant characteristics for the EFA and CFA samples. Participants in the EFA sample were generally younger than those in the CFA sample; half of the EFA sample was aged 15–24 years (n=977, 50%) compared with 31% (n=290) in the CFA sample. Across samples, most participants identified as White (EFA: n=1,456, 75%; CFA: n=697, 73%), reported at least some college education (EFA: n=1,351, 69%; CFA: n=717, 75%), and had an annual income of $75,000 or more (EFA: n=611, 36%; CFA: n=345, 40%). The largest proportion resided in the South (EFA: n=573, 29%; CFA: n=280, 29%). A small proportion of participants reported living with HIV (EFA: n=12, 1%; CFA: n=20, 3%).

**Table 1.** Participant demographics by sample.

|  | EFA Sample (n=1951) |  | CFA Sample (n=950) |  |
|  | n | % | n | % |
|---|---|---|---|---|
| Age |  |  |  |  |
| 15–24 | 977 | 50% | 290 | 31% |
| 25–29 | 432 | 22% | 202 | 21% |
| 30–39 | 379 | 19% | 262 | 28% |
| 40+ | 163 | 8% | 196 | 21% |
| Race |  |  |  |  |
| Black | 50 | 3% | 70 | 7% |
| Hispanic | 209 | 11% | 73 | 8% |
| White | 1456 | 75% | 697 | 73% |
| Other | 236 | 12% | 110 | 11% |
| Education |  |  |  |  |
| Less than high school | 136 | 7% | 49 | 5% |
| High school | 459 | 24% | 181 | 19% |
| Some college or technical degree | 678 | 35% | 358 | 38% |
| College or postgraduate | 673 | 35% | 359 | 38% |
| Income |  |  |  |  |
| \$0-19999 | 282 | 17% | 135 | 16% |
| \$20000-39999 | 388 | 23% | 175 | 20% |
| \$40000-74999 | 406 | 24% | 211 | 24% |
| \$75000+ | 611 | 36% | 345 | 40% |
| Region |  |  |  |  |
| Northeast | 333 | 17% | 140 | 15% |
| Midwest | 394 | 20% | 231 | 24% |
| South | 573 | 29% | 280 | 29% |
| West | 650 | 33% | 299 | 31% |
| U.S. Dependent Territories | 1 | 0% | 0 | 0% |
| HIV Status |  |  |  |  |
| Living with HIV | 12 | 1% | 20 | 3% |
| Not living with HIV | 1092 | 99% | 592 | 97% |

### Item endorsement

Table 2 and Figure 1 show the item endorsement for gender identity and sexual behavior stigma items for both samples. Across EFA and CFA samples, the most commonly endorsed gender identity stigma items were feeling scared to be in public places (85-91%) and feeling that family members made discriminatory remarks (78-79%). For sexual behavior stigma, the most frequently endorsed items across EFA and CFA samples were verbal harassment (36-38%) and concerns pertaining to future interactions with healthcare providers (32-40%)

**Figure 1.**
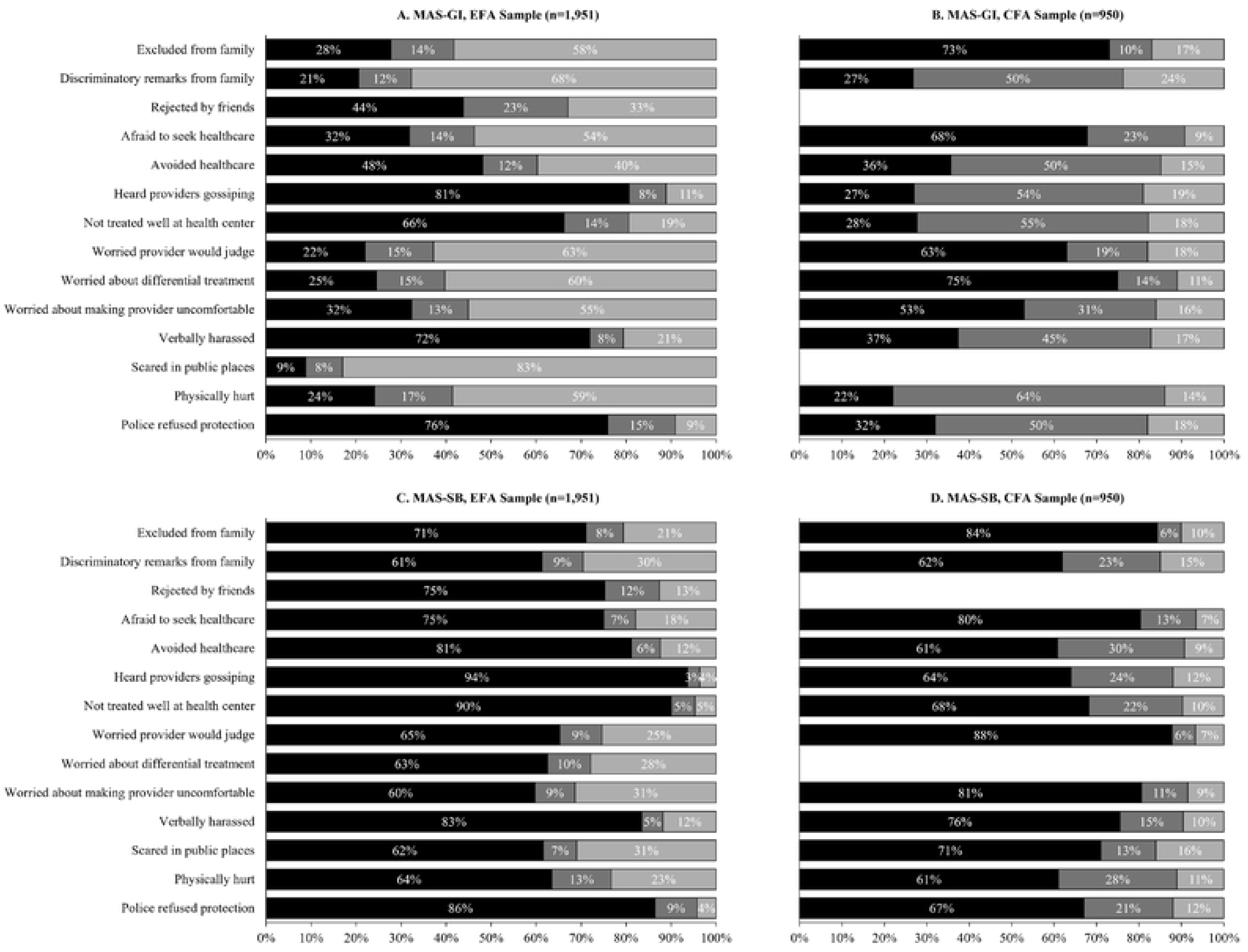
Item endorsement (%) for the Measures of Attributional Stigma scales by sample. MAS-GI = Measure of Attributional Stigma-Gender Identity; MAS-B = Measure of Attributional Stigma—Sexual Behavior, EFA = exploratory factor analysis; CFA = confirmatory factor analysis

**Table 2.**
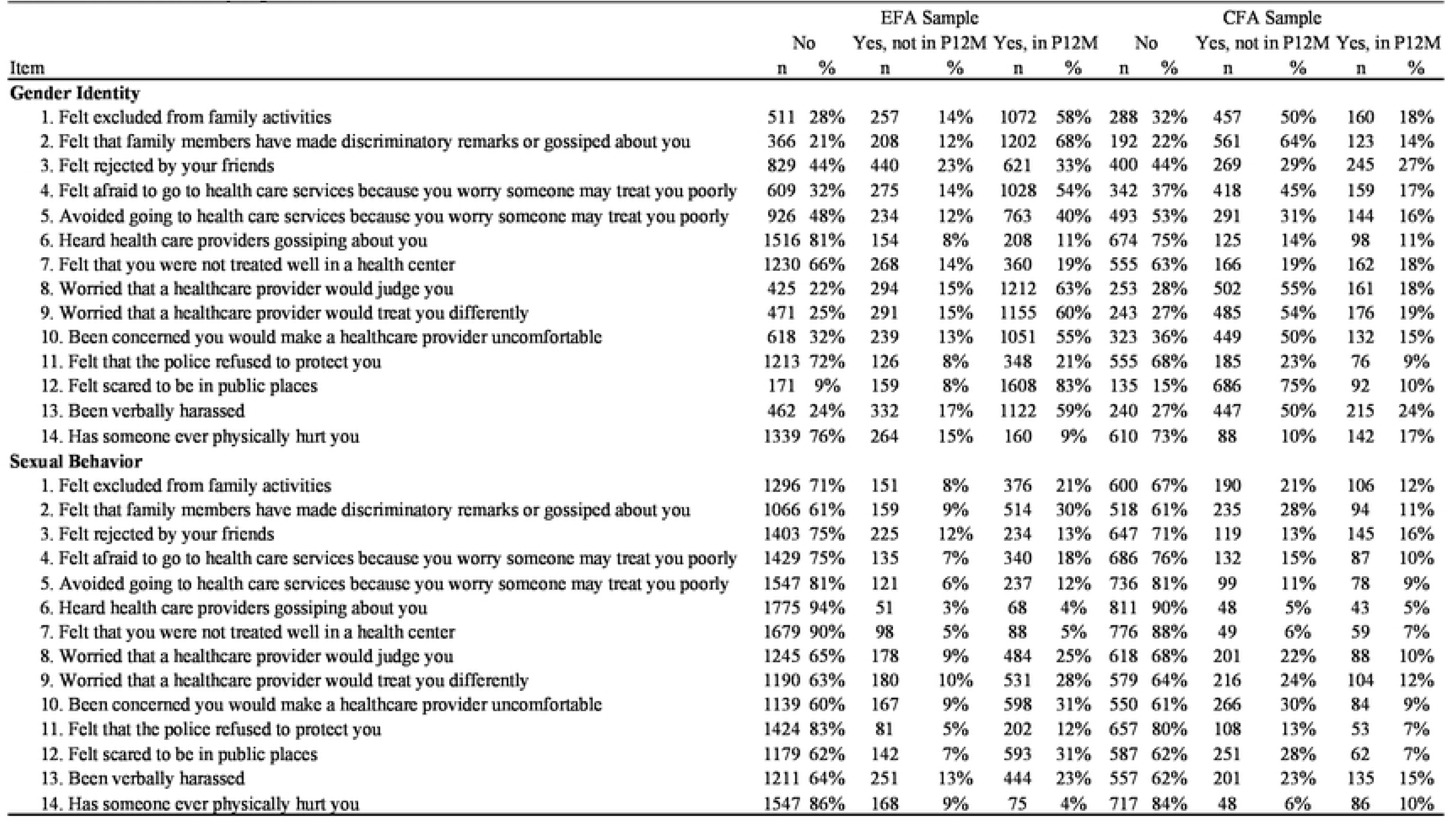
Gender and sexuality sligma items.

| Item | EFA Sample |  |  |  |  |  | CFA Sample |  |  |  |  |  |
|---|---|---|---|---|---|---|---|---|---|---|---|---|
|  | No |  | Yes, not in P12M |  | Yes, in P12M |  | No |  | Yes, not in P12M |  | Yes, in P12M |  |
| Item | n | % | n | % | n | % | n | % | n | % | n | % |
| **Gender Identity** |  |  |  |  |  |  |  |  |  |  |  |  |
| 1. Felt excluded from family activities | 511 | 28% | 257 | 14% | 1072 | 58% | 288 | 32% | 457 | 50% | 160 | 18% |
| 2. Felt that family members have made discriminatory remarks or gossiped about you | 366 | 21% | 208 | 12% | 1202 | 68% | 192 | 22% | 561 | 64% | 123 | 14% |
| 3. Felt rejected by your friends | 829 | 44% | 440 | 23% | 621 | 33% | 400 | 44% | 269 | 29% | 245 | 27% |
| 4. Felt afraid to go to health care services because you worry someone may treat you poorly | 609 | 32% | 275 | 14% | 1028 | 54% | 342 | 37% | 418 | 45% | 159 | 17% |
| 5. Avoided going to health care services because you worry someone may treat you poorly | 926 | 48% | 234 | 12% | 763 | 40% | 493 | 53% | 291 | 31% | 144 | 16% |
| 6. Heard health care providers gossiping about you | 1516 | 81% | 154 | 8% | 208 | 11% | 674 | 75% | 125 | 14% | 98 | 11% |
| 7. Felt that you were not treated well in a health center | 1230 | 66% | 268 | 14% | 360 | 19% | 555 | 63% | 166 | 19% | 162 | 18% |
| 8. Worried that a healthcare provider would judge you | 425 | 22% | 294 | 15% | 1212 | 63% | 253 | 28% | 502 | 55% | 161 | 18% |
| 9. Worried that a healthcare provider would treat you differently | 471 | 25% | 291 | 15% | 1155 | 60% | 243 | 27% | 485 | 54% | 176 | 19% |
| 10. Been concerned you would make a healthcare provider uncomfortable | 618 | 32% | 239 | 13% | 1051 | 55% | 323 | 36% | 449 | 50% | 132 | 15% |
| 11. Felt that the police refused to protect you | 1213 | 72% | 126 | 8% | 348 | 21% | 555 | 68% | 185 | 23% | 76 | 9% |
| 12. Felt scared to be in public places | 171 | 9% | 159 | 8% | 1608 | 83% | 135 | 15% | 686 | 75% | 92 | 10% |
| 13. Been verbally harassed | 462 | 24% | 332 | 17% | 1122 | 59% | 240 | 27% | 447 | 50% | 215 | 24% |
| 14. Has someone ever physically hurt you | 1339 | 76% | 264 | 15% | 160 | 9% | 610 | 73% | 88 | 10% | 142 | 17% |
| **Sexual Behavior** |  |  |  |  |  |  |  |  |  |  |  |  |
| 1. Felt excluded from family activities | 1296 | 71% | 151 | 8% | 376 | 21% | 600 | 67% | 190 | 21% | 106 | 12% |
| 2. Felt that family members have made discriminatory remarks or gossiped about you | 1066 | 61% | 159 | 9% | 514 | 30% | 518 | 61% | 235 | 28% | 94 | 11% |
| 3. Felt rejected by your friends | 1403 | 75% | 225 | 12% | 234 | 13% | 647 | 71% | 119 | 13% | 145 | 16% |
| 4. Felt afraid to go to health care services because you worry someone may treat you poorly | 1429 | 75% | 135 | 7% | 340 | 18% | 686 | 76% | 132 | 15% | 87 | 10% |
| 5. Avoided going to health care services because you worry someone may treat you poorly | 1547 | 81% | 121 | 6% | 237 | 12% | 736 | 81% | 99 | 11% | 78 | 9% |
| 6. Heard health care providers gossiping about you | 1775 | 94% | 51 | 3% | 68 | 4% | 811 | 90% | 48 | 5% | 43 | 5% |
| 7. Felt that you were not treated well in a health center | 1679 | 90% | 98 | 5% | 88 | 5% | 776 | 88% | 49 | 6% | 59 | 7% |
| 8. Worried that a healthcare provider would judge you | 1245 | 65% | 178 | 9% | 484 | 25% | 618 | 68% | 201 | 22% | 88 | 10% |
| 9. Worried that a healthcare provider would treat you differently | 1190 | 63% | 180 | 10% | 531 | 28% | 579 | 64% | 216 | 24% | 104 | 12% |
| 10. Been concerned you would make a healthcare provider uncomfortable | 1139 | 60% | 167 | 9% | 598 | 31% | 550 | 61% | 266 | 30% | 84 | 9% |
| 11. Felt that the police refused to protect you | 1424 | 83% | 81 | 5% | 202 | 12% | 657 | 80% | 108 | 13% | 53 | 7% |
| 12. Felt scared to be in public places | 1179 | 62% | 142 | 7% | 593 | 31% | 587 | 62% | 251 | 28% | 62 | 7% |
| 13. Been verbally harassed | 1211 | 64% | 251 | 13% | 444 | 23% | 557 | 62% | 201 | 23% | 135 | 15% |
| 14. Has someone ever physically hurt you | 1547 | 86% | 168 | 9% | 75 | 4% | 717 | 84% | 48 | 6% | 86 | 10% |

### Exploratory factor analysis

Sampling adequacy was high for both scales (overall KMO ≥0.85) and Bartlett’s tests of sphericity were significant (p<0.01) for both, supporting the suitability of the data for factor analysis. For the MAS-GI, three factors had eigenvalues exceeding 1 (5.57, 2.02, 1.23), and the parallel analysis and scree plot inspection also suggested a three-factor model (see Supplemental Table 1 and Supplemental Figure 1). Running the 3-factor solution and iteratively dropping poorly performing items (see Supplemental Table 2) resulted in Item 3 (“felt rejected by your friends”) and then item 12 (“felt scared to be in public places”) being dropped (both due to factor loadings <0.40). The final three factor solution cumulatively explained 73% of the variance.

Table 3 shows the final factor loadings and factor correlations for the MAS-GI. Factor 1 was made up of two items and reflected family stigma including perceived exclusion from family activities and discriminatory remarks from family. Factor 2 was made up of five items and reflected anticipated healthcare stigma including fear and avoidance of healthcare services and worries about healthcare provider treatment. Factor 3 was made up of five items and reflected enacted social stigma comprising experiences of mistreatment from healthcare providers and the police as well as being verbally harassed or physically attacked. Factor correlations were moderate between family stigma and anticipated healthcare stigma (r=0.30), family stigma and enacted social stigma (r=0.43), and anticipated healthcare stigma and enacted social stigma (r=0.42). Ordinal alpha was 0.86 for family stigma, 0.93 for anticipated healthcare stigma, and 0.83 for enacted social stigma.

**Table 3.** Factor loadings for gender stigma items (MAS-GI)

|  | EFA |  |  | CFA |  |  |
|---|---|---|---|---|---|---|
| Item | FAM | AHS | ES | FAM | AHS | ES |
| **Gender Identity** |  |  |  |  |  |  |
| 1. Felt excluded from family activities | **0.84** | 0.04 | 0.01 | 0.78 |  |  |
| 2. Felt that family members have made discriminatory remarks or gossiped about you | **0.86** | 0.02 | 0.01 | 0.75 |  |  |
| 4. Felt afraid to go to health care services because you worry someone may treat you poorly | 0.02 | **0.82** | 0.12 |  | 0.76 |  |
| 5. Avoided going to health care services because you worry someone may treat you poorly | 0.01 | **0.66** | 0.22 |  | 0.74 |  |
| 6. Heard health care providers gossiping about you | -0.02 | 0.18 | **0.69** |  |  | 0.80 |
| 7. Felt that you were not treated well in a health center | -0.07 | 0.28 | **0.67** |  |  | 0.82 |
| 8. Worried that a healthcare provider would judge you | 0.02 | **0.92** | 0.02 |  | 0.67 |  |
| 9. Worried that a healthcare provider would treat you differently | 0.02 | **0.94** | -0.10 |  | 0.82 |  |
| 10. Been concerned you would make a healthcare provider uncomfortable | 0.06 | **0.78** | -0.06 |  | 0.88 |  |
| 11. Felt that the police refused to protect you | 0.03 | -0.04 | **0.67** |  |  | 0.57 |
| 13. Been verbally harassed | 0.25 | -0.07 | **0.52** |  |  | 0.50 |
| 14. Has someone ever physically hurt you | 0.13 | -0.17 | **0.74** |  |  | 0.57 |
| **Factor Correlations** |  |  |  |  |  |  |
| Family Stigma | - | 0.30 | 0.43 | - | 0.44 | 0.61 |
| Anticipated Healthcare Stigma | 0.30 | - | 0.42 | 0.44 | - | 0.65 |
| Enacted Stigma | 0.43 | 0.42 | - | 0.61 | 0.65 | - |

*FAM* Family Stigma; *AHS* Anticipated Healthcare Stigma; *ES* Enacted Stigma

For the MAS-SB, two eigenvalues exceeded 1 (7.20, 1.35), and a two-factor solution was also suggested as optimal by the parallel analysis and scree plot inspection (see Supplemental Table 1 and Supplemental Figure 2). Running the 2-factor EFA resulted in the iterative dropping of Item 6 (“heard healthcare providers gossiping about you”) and then item 12 (“felt scared to be in public places”) due to cross-loadings (Supplemental Table 3). The final two factor solution cumulatively explained 71% of the variance.

Table 4 shows the final factor loadings and factor correlations for the MAS-SB. Factor 1 contained six items and reflected global healthcare stigma including both anticipation of stigma expressed as fear and avoidance of services and perceived/enacted experiences of stigma in healthcare settings, including hearing providers gossip about oneself or generally not being treated well. Factor 2 contained six items and reflected social/institutional stigma from family, friends, police, and unspecified actors. The correlation between global healthcare stigma and social/institutional stigma was moderate (r=0.66). Ordinal alpha was 0.95 for global healthcare stigma and 0.87 for social/institutional stigma.

**Table 4.** Factor loadings for sexual identity stigma items (MAS-SeBe)

|  | EFA |  | CFA |  |
|---|---|---|---|---|
| Item | HS | ICS | HS | ICS |
| **Sexual Identity** |  |  |  |  |
| 1. Felt excluded from family activities | -0.08 | **0.89** |  | 0.84 |
| 2. Felt that family members have made discriminatory remarks or gossiped about you | -0.01 | **0.85** |  | 0.80 |
| 3. Felt rejected by your friends | 0.11 | **0.61** |  | 0.79 |
| 4. Felt afraid to go to health care services because you worry someone may treat you poorly | **0.91** | 0.01 | 0.90 |  |
| 5. Avoided going to health care services because you worry someone may treat you poorly | **0.82** | 0.09 | 0.91 |  |
| 7. Felt that you were not treated well in a health center | **0.63** | 0.20 | 0.84 |  |
| 8. Worried that a healthcare provider would judge you | **0.98** | -0.06 | 0.91 |  |
| 9. Worried that a healthcare provider would treat you differently | **0.95** | -0.04 | 0.93 |  |
| 10. Been concerned you would make a healthcare provider uncomfortable | **0.82** | 0.01 | 0.84 |  |
| 11. Felt that the police refused to protect you | 0.26 | **0.40** |  | 0.75 |
| 13. Been verbally harassed | 0.26 | **0.51** |  | 0.73 |
| 14. Has someone ever physically hurt you | 0.09 | **0.64** |  | 0.72 |
| **Factor Correlations** |  |  |  |  |
| Healthcare Stigma | - | 0.66 | - | 0.78 |
| Interpersonal and Community Stigma | 0.66 | - | 0.78 | - |

*HS* Healthcare Stigma; *ICS* Interpersonal and Community Stigma

### Confirmatory factor analysis

Figure 2 shows the factor structure and CFA results for the MAS-GI and MAS-SB. For the MAS-GI, the hypothesized three-factor model demonstrated good model fit (CFI=0.980, TLI=0.980, RMSEA=0.062 [95% CI: 0.054–0.070], SRMR=0.065). Standardized factor loadings ranged from 0.67 to 0.82 for the anticipated healthcare stigma factor, 0.50 to 0.88 for the enacted social stigma factor, and 0.75 to 0.78 for the family stigma factor. Factor correlations were 0.44 between family stigma and anticipated healthcare stigma, 0.61 between family stigma and enacted social stigma, and 0.65 between anticipated healthcare stigma and enacted social stigma.

**Figure 2.**
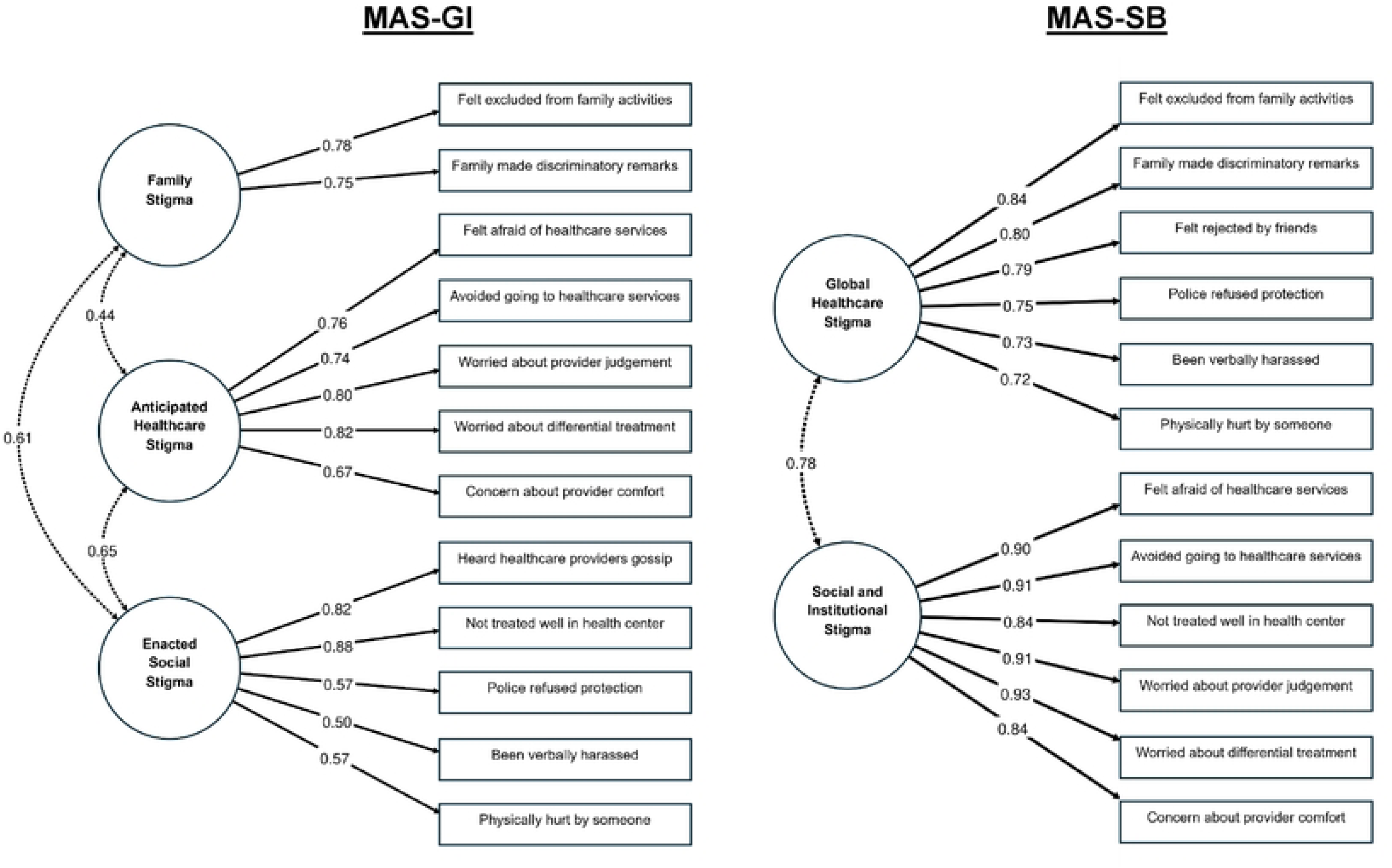
Confirmatory factor analysis models for the Measures of Attributional Stigma scales. MAS-GI = Measure of Attributional Stigma-Gender Identity; MAS-SB = Measure of Attributional Stigma—Sexual Behavior

For the MAS-SB, the hypothesized two-factor model also demonstrated excellent model fit (CFI=0.995, TLI=0.994, RMSEA=0.052 [95% CI: 0.044–0.060], SRMR=0.054). Standardized factor loadings were high for both factors, ranging from 0.84 to 0.93 for the global healthcare stigma factor and from 0.72 to 0.84 for the social/institutional stigma factor. Global healthcare stigma and social/institutional stigma had a factor correlation of 0.78.

### External construct validity

Table 5 shows the correlations and prevalence ratios between stigma subscales and psychosocial and healthcare utilization outcomes. For MAS-GI, higher levels of family stigma, anticipated healthcare stigma, and enacted social stigma were significantly positively correlated with depression symptoms (r=0.10-0.20) and posttraumatic stress symptoms (r=0.15-0.28). Anticipated healthcare stigma (r=-0.11) and enacted social stigma (r=-0.16) were significantly negatively correlated with social capital. Only anticipated healthcare stigma was significantly negatively associated with transgender pride (r=-0.10). In analyses of health service utilization, family stigma and enacted social stigma were associated with higher prevalence of HIV testing (PR=1.06-1.12) and STI testing (PR=1.09-1.18). Anticipated healthcare stigma was also associated with higher prevalence of HIV testing (PR=1.05). Enacted social stigma was also associated with greater lifetime PrEP use (PR=1.25). Family stigma was associated with a higher prevalence of mental health care utilization in the past 12 months (PR=1.09).

**Table 5.** Correlations and prevalence ratios between MAS-GI and MAS-3B subscales and external validation measures.

|  | Prevalence Ratios (95% Cis) |  |  |  |  | Correlations |  |  |  |
|---|---|---|---|---|---|---|---|---|---|
|  | HIV test (ever) | STI test (p12m) | PrEP use (ever) | See HCP (p12m) | See MHCP (p12m) | Depression | PTSD | Social Capital | Pride |
| **Gender Identity** |  |  |  |  |  |  |  |  |  |
| Family Stigma | **1.06 (1.02-1.11)** | **1.09 (1.00-1.19)** | 1.16 (1.00-1.34) | 1.02 (0.99-1.04) | **1.09 (1.04-1.15)** | **0.10** | **0.15** | -0.06 | 0.02 |
| Anticipated Healthcare Stigma | **1.05 (1.00-1.10)** | 0.97 (0.89-1.06) | 1.10 (0.95-1.28) | 1.00 (0.97-1.02) | 1.02 (0.97-1.07) | **0.20** | **0.18** | **-0.11** | **-0.10** |
| Enacted Stigma | **1.12 (1.07-1.16)** | **1.18 (1.09-1.27)** | **1.25 (1.10-1.41)** | 1.01 (0.99-1.04) | 1.04 (0.99-1.09) | **0.18** | **0.28** | **-0.16** | -0.02 |
| **Sexual Identity** |  |  |  |  |  |  |  |  |  |
| Healthcare Stigma | **1.07 (1.03-1.11)** | **1.13 (1.05-1.22)** | **1.27 (1.13-1.42)** | 1.01 (0.98-1.03) | **1.05 (1.01-1.10)** | **0.24** | **0.25** | **-0.12** | 0.01 |
| Interpersonal and Community Stigma | **1.06 (1.02-1.11)** | **1.16 (1.08-1.25)** | **1.29 (1.15-1.44)** | 0.98 (0.96-1.01) | 1.04 (1.00-1.09) | **0.19** | **0.28** | **-0.10** | 0.05 |

*Note* Bold values indicate p<0.05
*HCP* Healthcare Provider; *MHCP* Mental Health Care Provider

For the MAS-SB, both global healthcare stigma and social/institutional stigma were significantly correlated with depression (r=0.19-0.24) and posttraumatic stress symptoms (r=0.25-0.28) and negatively correlated with social capital (r=-0.10 to -0.12). Neither stigma domain was significantly correlated with transgender pride. In terms of health service utilization, both stigma domains were associated with higher prevalence of lifetime HIV testing (PR=1.06-1.07), STI testing in the past 12 months (PR=1.13-1.16), and lifetime PrEP use (PR=1.27-1.29). Global healthcare stigma was also associated with a higher prevalence of mental health care utilization in the past 12 months (PR=1.05).

## Discussion

The present study evaluated the factor structure and psychometric properties of the MAS-GI and MAS-SB scales among a large, geographically diverse sample of transgender women in the United States. Using EFA, we identified a three-factor structure for the MAS-GI comprising family stigma, anticipated healthcare stigma, and enacted social stigma and a two-factor structure for the MAS-SB comprising global healthcare stigma and social/institutional stigma. These factor structures were confirmed through CFA in an independent sample, demonstrating robust model fit. External validity analyses demonstrated consistent positive associations between all stigma subscales and depression and PTSD, while most stigma subscales were negatively associated with social capital. Findings for transgender identity pride and healthcare engagement outcomes were more mixed. These validated measures can support more precise measurement and surveillance of stigma across different sources and contexts, facilitate the identification of intervention targets, and inform evaluation of efforts to reduce stigma and its health consequences among transgender women.

The factor structures identified in the current study both extend and refine those reported by Wiginton, Maksut (22) where a two-factor structure for gender identity stigma (social stigma and institutional stigma/violence) and a single-factor structure for sexual behavior stigma were identified. In the prior gender-identity solution, family experiences were included with social stigma, whereas healthcare, police, and violence-related experiences formed an institutional stigma/violence factor. In the current study, the two family-related items formed a distinct family stigma factor; anticipated healthcare stigma formed a separate factor; and the remaining interpersonal, public, healthcare, and police-related experiences grouped into a general social stigma factor. Family rejection and acceptance may be particularly salient for transgender individuals given the documented importance of family support for transgender identity development and mental health (43, 44). The emergence of anticipated healthcare stigma as a separate factor also underscores healthcare-related concerns for this population, potentially reflecting the well-documented barriers and mistreatment transgender women face when seeking medical care (14, 45, 46).

For sexual behavior stigma, the prior one factor structure included family, friendship, healthcare, police, public, and interpersonal stigma experiences. In contrast, the present analysis distinguished healthcare stigma, encompassing both anticipated and enacted healthcare stigma, from global social stigma, which included family, peer, police, public, and interpersonal experiences. This finding suggests that healthcare may be a distinct context for stigma attributed to sexual behavior, particularly when care requires disclosure of sexual behaviors. Evidence on sexual behavior stigma among transgender women remains limited, with only a few studies linking this stigma to psychological distress (47, 48), and future research should examine how gender-identity and sexual-behavior attributions overlap across settings.

Sexual behavior stigma was less commonly endorsed than gender identity stigma. The MAS-SB measures whether participants attribute stigma experiences to sexual behavior, rather than directly assessing sexual orientation or particular sexual practices. Transgender women have diverse sexual orientations, relationships, bodies, gender expressions, and disclosure experiences and gender identity and sexual behavior stigma may overlap but become salient in different settings. Lower endorsement may reflect that participants more commonly attributed negative experiences to transgender identity or gender expression, rather than indicating that sexual behavior stigma is unimportant or absent. Gender nonconformity may be more visible in social interactions than sexual behavior, which is typically private (49). Additionally, the current sociopolitical climate has intensified the scrutiny of transgender identities specifically, potentially making gender identity more problematic within systems of cisheteronormativity and a more salient attribution for negative experiences.

Associations between stigma subscales and external correlates provided partial support for construct validity. All stigma dimensions demonstrated significant positive associations with depression and posttraumatic stress symptoms, consistent with minority stress theory, which posits that stigma and discrimination constitute chronic stressors that contribute to adverse mental health outcomes among marginalized populations (5-7). All stigma dimensions were also negatively associated with social capital. These factors likely have a reciprocal relationship where stigma can erode social connections and community ties, and social capital may act as a protection from both stigma and the effects of stigma (50-53). The association between anticipated healthcare stigma and lower transgender identity pride suggests that concerns about stigma in medical settings may uniquely undermine positive self-regard related to one’s gender identity, possibly because healthcare interactions often require disclosure and vulnerability around gender identity (54, 55). Additionally, the pervasive history and ongoing mistreatment of transgender individuals within medical institutions may make anticipated stigma in these settings particularly damaging to transgender identity pride. Other stigma dimensions were not significantly associated with pride, including all MAS-SB subscales which may reflect how the pride measure specifically assessed transgender identity. However, the absence of significant associations with other gender identity subscales requires further investigation, including consideration of whether additional items capturing other dimensions of stigma might show different patterns.

Several stigma subscales were positively associated with healthcare utilization outcomes including HIV testing, STI testing, PrEP use, and mental healthcare. The directionality is difficult to interpret in cross-sectional data as they may be related through multiple, potentially opposing pathways. Prior research indicates that stigma can discourage or delay HIV testing, engagement in care, and PrEP uptake among transgender individuals (56-58). Conversely, those who are more engaged with healthcare may have more opportunities to experience stigma. Transgender women who access healthcare services may encounter stigma during these interactions, particularly when sexual health services require disclosure of behaviors that may elicit provider judgment (54, 59). Additionally, individuals with greater healthcare needs (potentially driven by stigma-related health consequences) may have more contact with the healthcare system (16, 60). Longitudinal research is needed to examine the temporal ordering and possibly bidirectional nature of this relationship.

The validated measures in this study have multiple research and practice applications. The MAS scales allow researchers to characterize the nature and magnitude of stigma, directly compare stigma attributed to gender identity versus sexual behavior, and identify subgroups at greatest risk. The emergence of distinct factors related to the source of stigma points to specific domains that may warrant targeted assessment and intervention. These scales can allow researchers to measure stigma with greater specificity, allowing a better understanding of the ways in which people attribute stigma from different sources to certain identities or attributes and how these may interact to shape key health outcomes across various populations and contexts. Furthermore, the scale design allows for the attributions to be easily adapted for a variety of stigmatized identities, allowing for the study of a broader range of populations and the different ways they may be affected by stigma.

Several limitations should be considered when interpreting these findings. The sample was recruited through online convenience sampling and is not representative of all transgender women. The EFA and CFA samples differed substantially in age composition, with the CFA sample being notably older. While the confirmation of factor structures across these samples strengthens confidence in the robustness of the measures, age-related differences in stigma experiences should be considered when interpreting findings. Older participants may have accumulated more lifetime stigma experiences and cohort effects may influence both exposure to and reporting of stigma given evolving social attitudes toward transgender individuals (e.g., older cohorts may have come of age with fewer legal protections and greater pathologization of transgender identity). Future research should formally evaluate measurement invariance across age groups to confirm that the scales function equivalently across the lifespan. The high correlation between the MAS-SB global healthcare stigma and social/institutional stigma factors indicates substantial overlap and warrants replication and further assessment of their discriminant validity. The stigma items relied on self-report and participants’ attributions of negative experiences to gender identity or sexual behavior, which may be subject to bias, and some experiences may be difficult to attribute definitively when identities or behaviors intersect. Finally, the measures assessed primarily anticipated, perceived, and enacted stigma; internalized stigma was not assessed and is conceptually distinct, warranting separate development and assessment of measurement.

## Conclusions

Stigma remains a pervasive and damaging force in the lives of transgender women. Rigorous measurement of stigma and its consequences among transgender populations has never been more urgent. The measures explored in this study equip researchers and practitioners to assess experiences of stigma based on cisnormativity, manifesting in gender identity stigma, and heteronormativity, manifesting in sexual behavior stigma, with greater precision, distinguishing both by stigma mechanism (e.g., enacted, anticipated) and the context in which each occurs (e.g., healthcare, family, community). These scales can be used to document the burden of stigma in its different forms, inform targeted interventions for transgender women, and evaluate whether efforts to reduce specific forms of stigma are successful.

## Acknowledgements

We gratefully acknowledge the transgender women who participated in this study and shared their experiences.

## Data Availability Statement

The data that supports the findings of this study are available from the corresponding author upon request.

